# Fear among Syrians: a Proposed Cutoff Score and Validation of the Arabic Fear of COVID-19 Scale- A National Survey

**DOI:** 10.1101/2021.05.25.21257637

**Authors:** Fatema Mohsen, Batoul Bakkar, Salma Khadem Alsrouji, Esraa Abbas, Alma Najjar, Marah Marrawi, Youssef Latifeh

## Abstract

COVID-19 pandemic has led to psychological health issues one of which is fear. This study validates the Arabic version of the fear of COVID-19 scale and suggests a new cutoff score to measure fear of COVID-19 among the Syrian Population. A total of 3989 participants filled an online survey consisting of socio-demographic information, the fear of COVID-19 scale, the patient health questionnaire 9-item, and the generalized anxiety disorder 7-item. Receiver operating characteristic analysis was used to define cutoff scores for the fear of COVID-19 scale in relation to generalized anxiety disorder 7-item and the patient health questionnaire 9-item. The Cronbach α value of the Arabic fear of COVID-19 scale was 0.896, revealing good stability and internal consistency. The inter-item correlations were between [0.420 – 0.868] and the corrected item-total correlations were between [0.614 – 0.768]. A cutoff point of 17.5 was deduced from analysis. According to the deduced cutoff point, 2111(52.9%) were classified as cases with extreme fear. This cutoff score deduced from this study can be used for screening purposes to identify individuals that may be most vulnerable towards developing extreme fear of COVID-19. Therefore, early preventive and supportive measures can then be delivered.

## Introduction

The novel coronavirus disease-2019 (COVID-19) is a serious threat to humanity’s health that has emerged as an outbreak and was declared by the World Health Organization (WHO) as a Public Health Emergency of International Concern (PHEIC).(1) The first incident was noted in Wuhan, China in December 2019. This virus quickly crossed the borders, spreading all over the world, resulting in a never-ending pandemic burdened with the effects of morbidity and mortality.(2) There are live updates of new cases, and deaths every second of the day, as well as official briefings, and prognostications about future calamity.(3) Enumeration of all deaths, when compared to the 0.28% crude mortality rate, which will continue to rise as more infections and deaths occur, produces a picture of excess death, capturing both the direct burden of the pandemic and its indirect mortality burden.(4, 5) 22 March 2020 marked the first officially reported COVID-19 case in Syria.(6) The numbers have escalated since then with Syria now entering its third wave.(7-9)

As the pandemic has embedded misconceptions around COVID-19, strict precautionary measures have been adopted by governments, such as physical distancing, self-isolation, and handwashing(10) to curtail the tide of the pandemic in the absence of vaccine or treatment at the time of the survey. During the Syrian lockdown, many streets have become ghost towns. Flights to and from affected parts of the world have been grounded. Business conferences have been cancelled. Mosques, museums, parks, schools, and universities have been closed and abandoned. Grocery and drug store shelves are being emptied of cold and flu remedies, hand sanitizer, disinfecting wipes, and anti-inflammatory over-the-counter medications. People are stacking their shelves with food, drinks, and toiletries, preparing for a long quarantine period that may never come to an end. The disruption in normal daily routines with social distancing and self-quarantine will cause lasting economic consequences given supply and demand-side shocks. With businesses closed and people avoiding public places, less money and fewer goods and services are exchanging hands. The effects of the media have had a toll on society, the reaction by the media and government is likely to produce more harm to societies around the globe than the virus, possibly for many years to come. As a major health issue, the COVID-19 pandemic has triggered a multilevel global crisis, affecting individual’s physical and mental health. Thus, researchers are expressing concerns related to COVID-19 adverse effects on society’s mental health and psychological well-being.(11-13)

One of these psychological impacts is fear, an important emotion for an individual’s survival by arousing adaptive defense responses against potentially threatening events or danger.(14) The current threatening event is COVID-19, a negative emotional reaction to or persistent worry over contracting the virus, illness, and death.(15) Added to the previous are changes in our daily lives working from home, temporary unemployment, home-schooling, and lack of socializing. The fear is likely to become a more pressing concern, as the pandemic sustains high infection and mortality rates. These concerns have pushed researchers to assess the fear of COVID-19 in different regions of the world and identify the possible triggers.(16)

Fear may lead to the incidence of new mental health disorders or worsening pre-existing psychiatric symptoms such as depression, anxiety, and post-traumatic stress disorder, and even suicide.(13, 17, 18) A recent study has developed the Fear of COVID-19 Scale (FCV-19S) to complement the clinical efforts in preventing the spread and treating of COVID-19 cases.(19)A few studies thereafter have validated the questionnaire in various languages and examined associations with socio-demographic characteristics.(20-27) However, only one study as of yet has proposed a cutoff score for the scale.(28) Given that Syria is one of the most vulnerable countries in the world to be affected by mental health disorders due to both COVID-19 and war. This study aimed to assess the fear of COVID-19 among the Syrian population, validate the FCV-19S, and identify an appropriate cutoff score to differentiate adults with extreme fear of COVID-19 from those with a normal fear reaction.

## Materials and methods

### Study design, setting, and participants

This web-based cross-sectional study was conducted using an Arabic questionnaire over a period of 12 days between May 2 and May 14 of 2020. The sample size calculated was 2401 participants based on a margin of error of 2%, and a confidence interval of 95%, for a population of 17, 500, 657 people using a sample size calculator.(29, 30) All participants aged 18 and above, residing in Syria, who completed the survey, were included in the study. The questionnaire was distributed through various social media platforms. After providing informed consent online, participants were directed to the first part of the survey to complete questions about socio-demographic information including gender, age, residence, education level, occupation, and economic status. Participants were also asked about the history of chronic diseases. To validate the Arabic version of the FCV-19S, a dataset comprising 3989 participants was used

### Adaptation of FCV-19S into Arabic

The original FCV-19S was translated into Arabic using a forward-backward translation technique.(19) First, the FCV-19S was translated into Arabic by two medical translators who are fluent in both Arabic and English. The authors, who are fluent in Arabic and English, reviewed the provisional Arabic translation and no anomalies were identified with the translation. Third, the approved Arabic translation draft of the scale was then back translated into English by another medical translator who was unfamiliar with the original English FCV-19S. Both the forward and backward translations of the scale were then compared for equivalence and checked for cultural appropriateness among the authors. The approved Arabic questionnaire was piloted with 20 volunteers to assess the scale’s reliability, clarity, relevance, and acceptability of the survey. These volunteers were excluded from the final sample and statistical analysis to avoid bias. Based on the feedback no modifications were required. Both the Arabic and English versions of the FCV-19S are provided in S1 appendix.

### Measures

The FCV-19S was used to evaluate the symptoms of fear. For this study, we used the Arabic version of FCV-19S, which was validated in a previous study.(31) The FCV-19S is a one-dimensional scale that measures one’s fear level of COVID-19 it consists of 7 items and is scored on a 5-point scale, ranging from 1 (strongly disagree) to 5 (strongly agree). A total score is calculated by summing all item scores with a possible total score ranging between 7 and 35. Higher scores indicate greater levels of fear of COVID-19.(26)

The Arabic version of the Patient Health Questionnaire 9-item (PHQ-9) was used in the study to assess depression symptom severity.(32) Items on the PHQ-9 were rated on a 4-point Likert scale (0=not at all, 1=several days, 2=more than half the days, 3=nearly every day), providing a 0-27 severity score range. The scores were categorized into 5 groups: none (0–4), mild (5–9), moderate (10–14), moderately severe (15-19), and severe (20–27).

The Arabic version of the Generalized Anxiety Disorder 7-item (GAD-7) was used in this study to assess anxiety symptom severity.(33) Items on the GAD-7 were rated on a 4-point Likert scale (0=not at all, 1=several days, 2=more than half the days, 3=nearly every day), providing a 0-21 severity score range. The Gad-7 is a self-rated scale used to evaluate the severity of the 4 most common anxiety disorders (Generalized Anxiety Disorder, Panic Disorder, Social Phobia, and Post Traumatic Stress Disorder), where a cutoff score of 0–4 indicates no anxiety symptoms. The scores were categorized into 4 groups: none (0-4), mild (5-9), moderate (10-14), and severe (15-21).(34)

### Ethics

The study was ethically approved by the Institutional Review Board (IRB) of the Faculty of Medicine, Syrian Private University.

### Statistical analysis

Data analysis was conducted using the Statistical Package for Social Sciences version 25.0 (SPSS Inc., Chicago, IL, United States). Descriptive analysis, including frequencies, percentages, means, and standard deviations (SD) were applied. The Cronbach’s α test and inter-item correlation were used to assess the internal consistency, with satisfactory reliability set at ≥ 0.70 and between 0.20 and 0.40 respectively.(35, 36) The corrected item-total correlation was used to assess the coherence of the FCV-19S (values > 0.4 are acceptable). The current analysis focuses on the association between the FCV-19S scale and other variables relevant to the participants’ psychological reaction to COVID-19, including anxiety, and depression. Receiver operating characteristic (ROC) analysis was used to define cutoff scores for the FCV-19S scale in relation to GAD-7 and PHQ-9 serving as external criteria. The Youden-Index was used to determine the optimal cutoff score and to reduce the risk of misclassification. A dichotomous variable was created out of the total GAD-7 using the cutoff point of 10 to assess anxiety symptoms.(34) A dichotomous variable was created out of the total PHQ-9 using the cutoff point of 10 to assess depressive symptoms.(32) After identifying the cutoff points, participants with a sum-score above the given cutoff value were considered cases with extreme fear. Those below were regarded as non-fear cases.

## Results

### Socio-demographic characteristics of participants

Of 5000 total participants invited to take part in the study, 4,430 gave informed consent. A final sample size of 3989 participants (response rate = 79.8%) met the inclusion criteria for the study. Most participants were female 2935 (73.5%), single 3096 (77.6%), students 2397 (60.1%), and residing in Damascus 1412 (35.4%). Participant ages ranged from 18 to 70 years, with the age group 18-25 years representing a majority 2870 (71.9%). A total of 416 (10.4%) and 1522 (38.1%) participants stated they had poor and moderate economic status respectively. 556 (15.9%) mentioned a history of chronic diseases Table 1.

**Table 1.** Socio-demographic characteristics: (n=3989)

|  |  |  |  |  |  |
| --- | --- | --- | --- | --- | --- |
| **Gender(%)** | **Male** | **1054(26.4%)** | **Education (%)** | **Primary School** | **21(0.5%)** |
|  | **Female** | **2935(73.5%)** |  | **Intermediate School** | **115(2.9%)** |
| **Age(%)** | **18-25** | **2870(71.9%)** |  | **Secondary school** | **370(9.3%)** |
|  | **26-34** | **685(17.2%)** |  | **College/University** | **3271(82%)** |
|  | **35-44** | **261(6.5%)** |  | **Master's degree** | **185(4.6%)** |
|  | **45-54** | **121(3%)** |  | **PhD** | **27(0.7%)** |
|  | **55 <** | **52(1.4%)** | **Occupation(%)** | **Health care worker** | **259(6.5%)** |
| **Social Status(%)** | **Single** | **3096(77.6%)** |  | **Government institution** | **239(6%)** |
|  | **Married** | **714(17.9%)** |  | **Private institution** | **202(5.1%)** |
|  | **Other** | **179(4.5%)** |  | **Business** | **202(5.1%)** |
| **Economic** | **Excellent** | **251(6.3%)** |  | **Military** | **35(0.9%)** |
| **Status(%)** | **Good** | **1800(45.1%)** |  | **Student** | **2397(60.1%)** |
|  | **Moderate** | **1522(38.1%)** |  | **Other** | **655(16.4%)** |
|  | **Poor** | **416(10.4%)** | **Household members(%)** | **Alone** | **54(1.4%)** |
| **Chronic disease(s)(%)** | **Yes** | **556(13.9%)** |  | **1-5** | **2474(62%)** |
|  | **No** | **3433(86%)** |  | **>5** | **1461(36.6%)** |

### Validity and Cutoff Score analysis

The FCV-19S scores ranged between 7 and 35, the mode score was 14, and the mean score was 18.5 (± 6.009) Fig 1. Participants’ responses to the FCV-19S are shown in Table 2. The Cronbach α value of the Arabic FCV-19S was 0.896, revealing good stability and internal consistency. The inter-item correlations were between [0.420 – 0.868] and the corrected item-total correlations were between [0.614 – 0.768].

**Table 2.** Participants response to FCV-19S.

| **Table 2. Participants response to FCV-19S** |  |  |  |  |  |
| --- | --- | --- | --- | --- | --- |
|  | **Totally disagree(%)** | **Disagree(%)** | **Neutral(%)** | **Agree(%)** | **Totally Agree(%)** |
| **1-I am often afraid of the emerging COVID-19** | (6.5)261 | 826(20.7) | 1557(39.0) | 951(23.8) | 394(9.9) |
| **2- I feel uncomfortable when I think about the new COVID-19** | (7.0)279 | 906(22.7) | 1115(28.0) | 1228(30.8) | 461(11.6) |
| **3- My hands get sweaty when I think about the new COVID-19** | (24.1)962 | 1983(49.7) | 719(18.0) | 119(3.0) | 206(5.2) |
| **4- I am afraid of dying due to infection with the emerging COVID-19** | (17.2)685 | 1579(39.6) | 892(22.4) | 463(11.6) | 370(9.3) |
| **5- I feel anxious and nervous when I follow news or posts about COVID-19 on social media** | 390(9.8) | 968(24.3) | 1002(25.1) | 1070(26.8) | 559(14.0) |
| **6- I cannot sleep because of my anxiety about infection with the emerging COVID-19** | 1045(26.2) | 1875(47.0) | 688(17.2) | 152(3.8) | 229(5.7) |
| **7- I feel my heart racing and palpitations when I think about the emerging COVID-19** | **1023(25.6)** | **1843(46.2)** | **670(16.8)** | **211(5.3)** | **242(6.1)** |

The responses to the GAD-7 scale were classified into two groups: non-anxious symptoms (score < 9) and anxious symptoms (9 < score ≤ 21). The responses to the PHQ-9 scale were classified into two groups: non-depressed symptoms (score < 9) and depressed symptoms (9 < score ≤ 27) Table 3.

**Table 3.** Classification of participants according to FCV-19S, GAD-7, and PHQ-9 cutoff points.

|  |  |  |
| --- | --- | --- |
|  | **Yes(%)** | **No(%)** |
| **FCV-19S** | 211(52.9) | 1878(47.1) |
| **GAD-7** | 1484(37.2) | 2505(62.8) |
| **PHQ-9** | 2176(54.6) | 1813(45.4) |

ROC analysis was used to evaluate the efficacy of the FCV-19S in predicting anxiety and depression factors. The proposed cutoff score was determined by the optimal sensitivity and specificity level, Table 4, revealing the following:

**Table 4.** prediction validity of the FCV-19S.

| **Table 4. prediction validity of the FCV-19S** |  |  |  |  |  |  |  |
| --- | --- | --- | --- | --- | --- | --- | --- |
|  | **FCV-19S** | **Sensitivity** | **Specificity** | **AUC** | **95%CI** | **p-value** | **Youden’s Index** |
| **Anxiety** | **18.5** | **0.532** | **0.575** | **0.560** | **[0.541 , 0.578]** | **0.001>** | **0.107** |
| **Depression** | **17.5** | **0.551** | **0.496** | **0.520** | **[0.502 , 0.538]** | **0.001>** | **0.047** |

- Unsatisfactory predictive power for the FCV-19S in disclosure of anxiety symptoms assessed by GAD-7 was found. The best cutoff was at 18.5, with a sensitivity of 53% and specificity of 57.5%, as the area under the curve (AUC) was 0.560 with a statistical significance (p-value < 0.001), a confidence interval of 95% CI=[0.541, 0.578] Table 4 and Fig 2.
- Unsatisfactory predictive power for the FCV-19S in disclosure of depressive symptoms assessed by PHQ-9 was found. The best cutoff was at 17.5 with a sensitivity of 55.1% and specificity of 49.6%, AUC=0.520 was statistically significant (p-value < 0.001), a confidence interval of 95% CI= [0.502, 0.538] Table 4 and Fig 3.

A cutoff point of 17.5 was deduced from the anxiety and depression ROC analysis. According to the deduced cutoff point, the majority 2111(52.9%) were classified as cases with extreme fear Table 3.

## Discussion

The current study was performed to validate the Arabic version of the FCV-19S and propose a satisfying cutoff to differentiate those with extreme fear from those with normal fear reactions towards COVID-19. Obtaining a cutoff is a pivotal and common practice in the psychiatry field to differentiate those into either cases or non-cases.(37-41) Identifying cases and non-cases with extreme fear will allow us to stratify those in need for further assessment by a specialist in psychiatry. Once assessed, appropriate management can be given if indicated. Although many studies have used the FCV-19S to assess fear among various populations, without a cutoff score we cannot further analyze the results and compare and interpret findings with different populations.(42)

Our study revealed a good internal consistency (Cronbach’s α = 0.896). Our finding was in line with other studies with a range of 0.82 to 0.88.((19, 43, 44) Saudi Arabian study(45) The inter-item correlations were between [0.420 – 0.868] and the corrected item-total correlations were between [0.614 – 0.768], whereas it was [0.35 - 0.66] and [0.57-0.74] respectively in a Saudi-Arabian study.(45)

ROC curves were constructed to determine the FCV-19S cutoff point. A cutoff point of 17.5 was deduced, participants scoring ≥ 17.5 were classified as having extreme fear of COVID-19, whereas participants scoring below this threshold were classified as having normal fear of COVID-19. Only one study has defined a cutoff for the FCV-19S, with a threshold of 16.5 and higher.(28)

This study revealed a weak discriminatory ability from the AUC outcomes, whereas its accuracy was moderate in a Greek study(46). The low precision of AUC signifies that not all individuals with depression or anxiety symptoms have an unhealthy fear of COVID-19 and vice versa. This may be attributed to ten years of conflict and the implications it induced on the mental health of the Syrian population (47). Thus, Syrians have countless factors contributing to their depressive and anxiety symptoms unlike the majority of the globe. Also, the FCV-19S was designed to assess fear towards COVID-19 while the GAD-7 and PHQ-9 are not aimed at assessing depression and anxiety towards the COVID-19 pandemic.

Our study revealed a prevalence of 52.9% of Syrians were classified as having extreme fear towards COVID-19. The prevalence of extreme fear among Syrians was higher in comparison with the Greeks (40.3%), despite their lower cutoff. The higher prevalence among Syrians can be attributed to the devastating impacts of war, driving the country into a prolonged economic recession. The devastating conflict has resulted in a Syrian healthcare system that is severely under-equipped and lacks the capacity to contain such a pandemic. With many healthcare professionals fleeing the country, a low number of ventilators, and many hospitals bombarded to rubble, Syrians must feel constant fear for their lives. (48)

All the above factors have exacted immense effects on the mental health of Syrians. The Syrian ministry of health provides three hospitals for mental illness and substance abuse: Ibn Rushd Hospital (Damascus), Ibn Sina Hospital (Rural Damascus), and Ibn Khaldoun Hospital (Aleppo). On 25 December 2012, Ibn Khaldoun Hospital was bombed, leaving only 2 hospitals standing. Our study showed that 54.6% and 37.2% of Syrians are suffering from moderate to severe depressive and anxiety symptoms respectively. Grievously, 90% of cases are left unattended, untreated, and unmanaged, as only 80 psychiatrists are working in Syrian territories.(49, 50) We must call on national and international organizations to aid us Syrians, reconstruct mental health services, and assist in providing skilled healthcare workers for the suffering people of Syria.

## Limitations

The present study has several limitations in its design. First, the convenience sampling used in this study may have limited results’ generalizability due to a “volunteer-effect” and the potential underrepresentation of less educated and socially disadvantaged participants. Second, credible published national data regarding the socio-demographic characteristics of Syrians are not available to evaluate the representativeness of our sample. Third, the cross-sectional design limited the opportunity to study the prospective effects of fear over time. Fourth, the proposed cutoff aimed to further evaluate the scale’s predicting ability to screen for cases with extreme fear, and not for diagnostic purposes. Fifth, a lack of other instruments assessing the same construct during the conduct of the study such as health anxiety and posttraumatic stress disorder. Sixth, the cutoff should ideally only be used on Syrian populations due to the characteristics of the participants in the study. Therefore, the proposed cutoff point must be interpreted with particular caution.

## Conclusion

This study provided empirical support for the scale’s adequacy to assess COVID-19-related fear and determined a cutoff point of ≥ 17.5 with unsatisfactory predictive power for anxiety and depression. This cutoff score can be used for screening purposes to identify individuals that may be most vulnerable towards developing psychiatric symptomatology. Therefore, require further assessment to identify high-risk individuals and deliver early preventive and/or supportive measures. However, the Syrian healthcare system is under constant strain due to the effects of war and the COVID-19 pandemic, and such measures may only exist in our imaginations.

## Data Availability

All data related to this paper are available within the manuscript.

## Acknowledgments

We are thankful to the management of the Syrian Private University for the support in the field of medical training and research. We are thankful to everyone who participated in this study.

## Abbreviations

COVID-19: Coronavirus Disease 2019
WHO: World Health Organization
PHEIC: Public Health Emergency of International Concern
FCV-19S: Fear of COVID-19 Scale
PHQ-9: Patient Health Questionnaire 9-item
GAD-7: Generalized Anxiety Disorder 7-item
ROC: Receiver operating characteristic
IRB: Institutional Review Board
SPSS: Statistical Package for Social Sciences
SD: Standard Deviation
AUC: Area Under the Curve

## Attached Survey

## Appendix 1

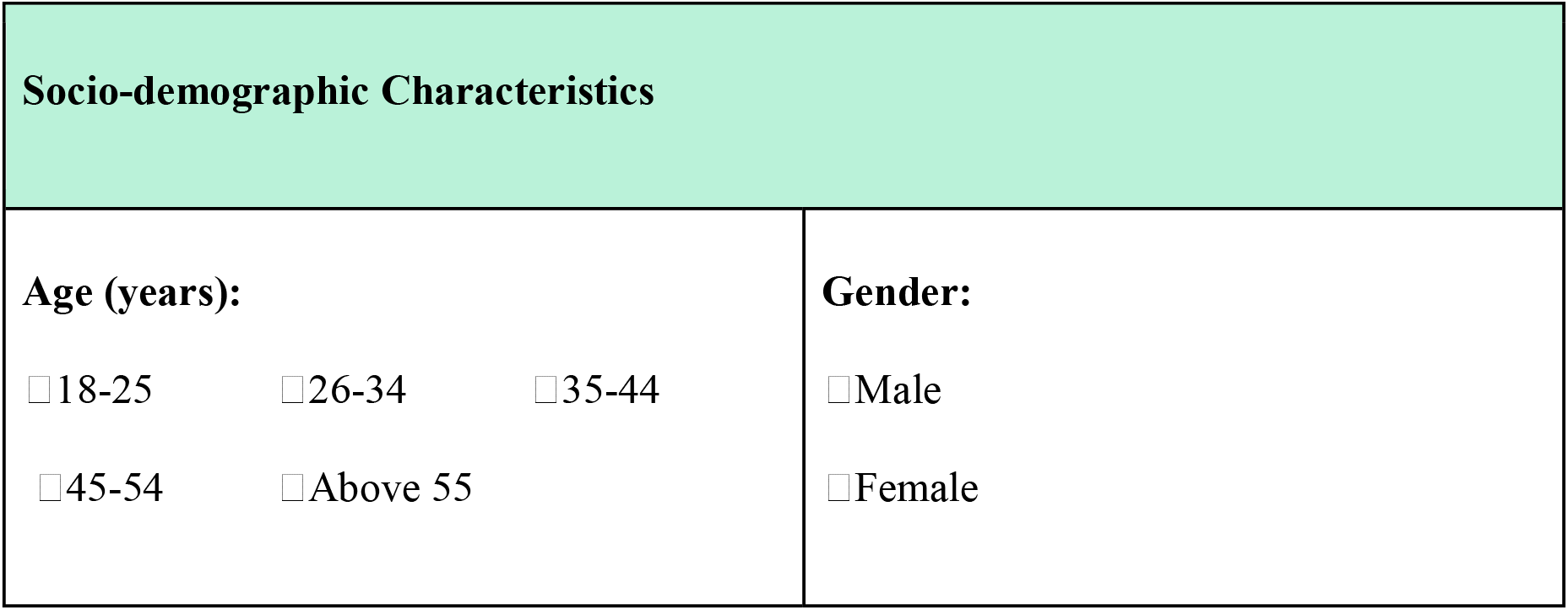
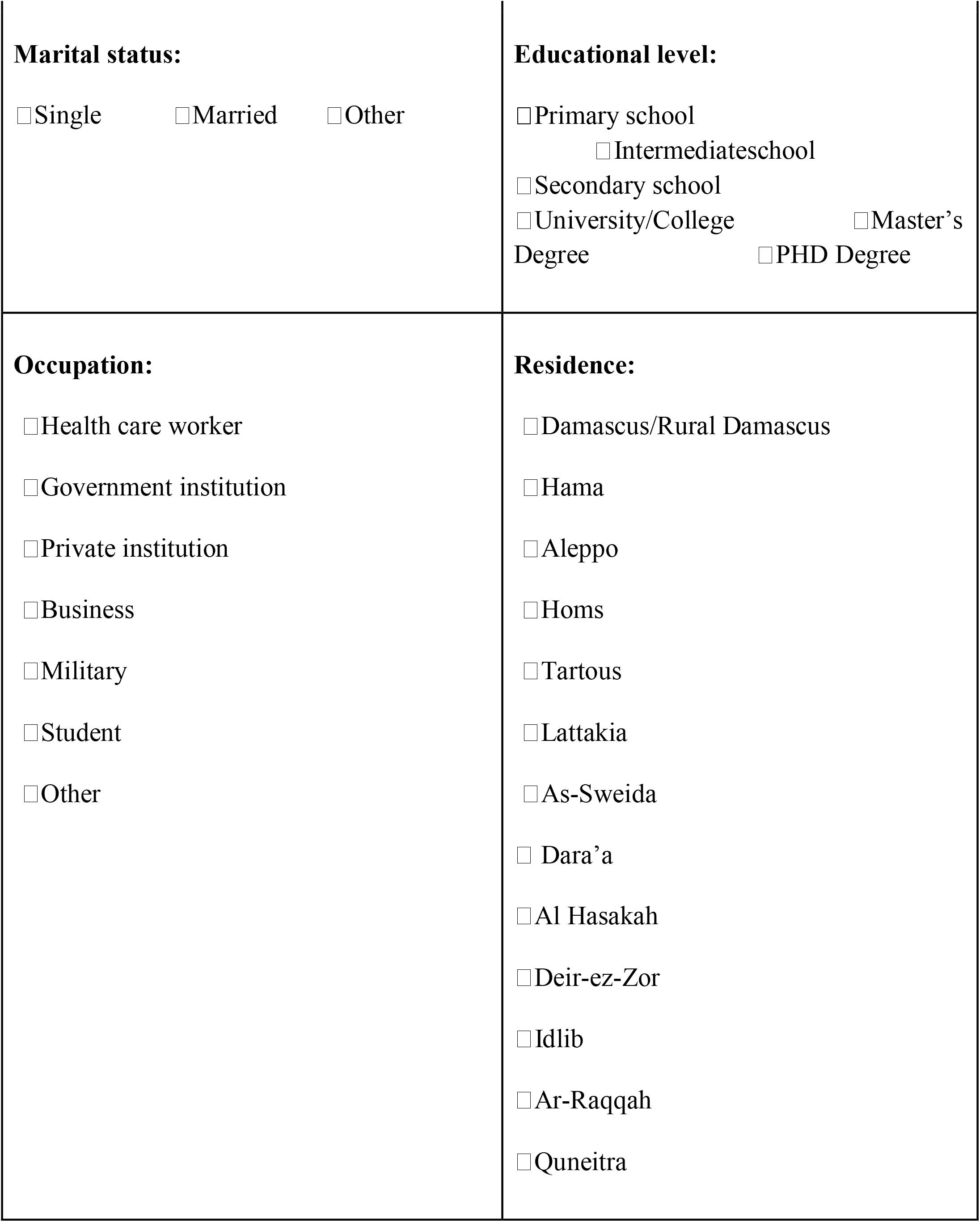
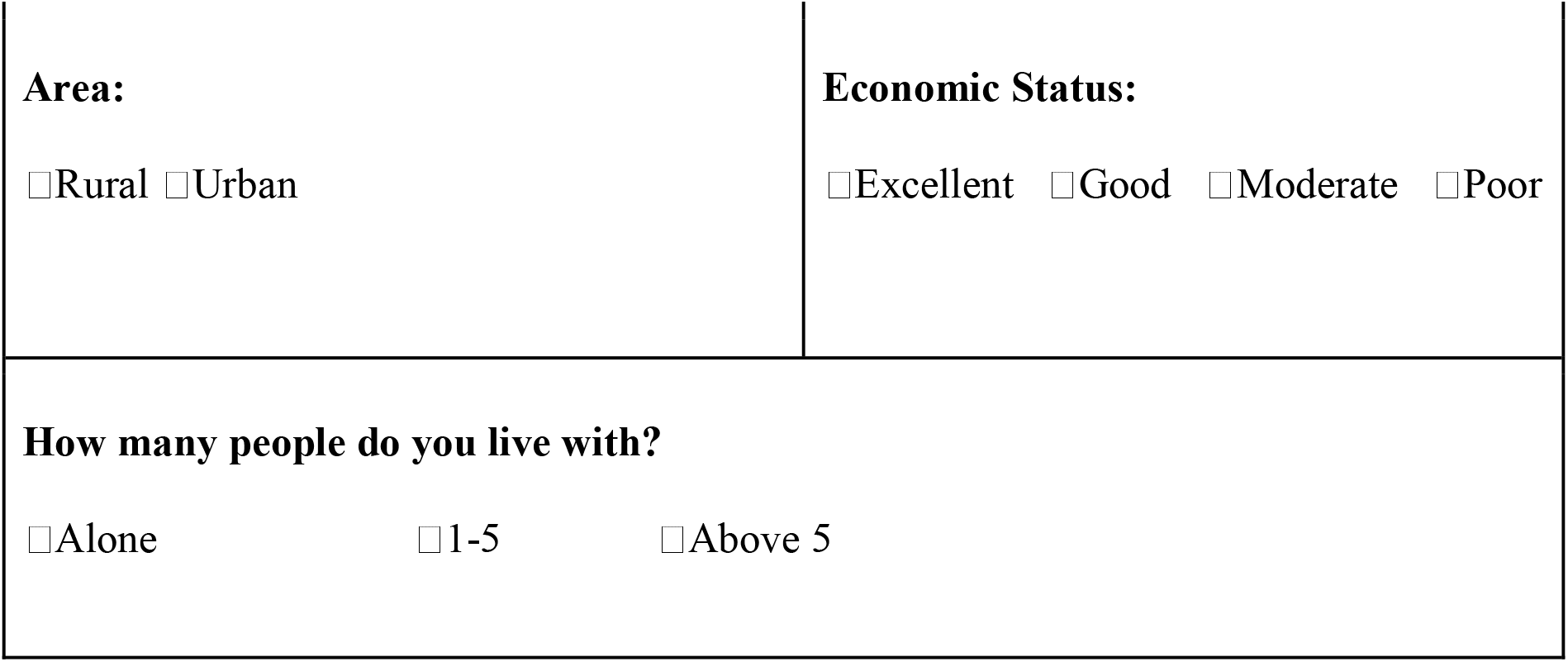

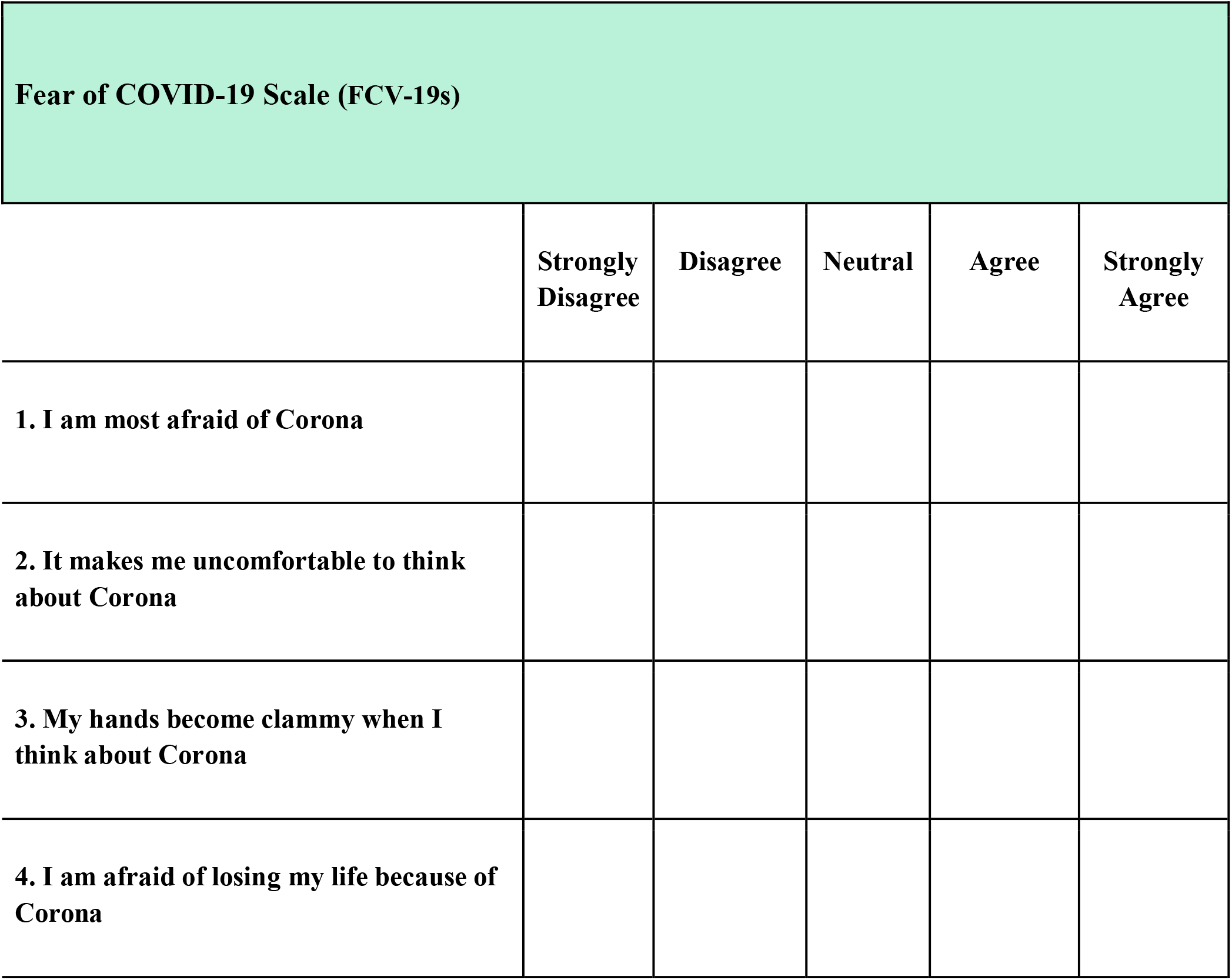
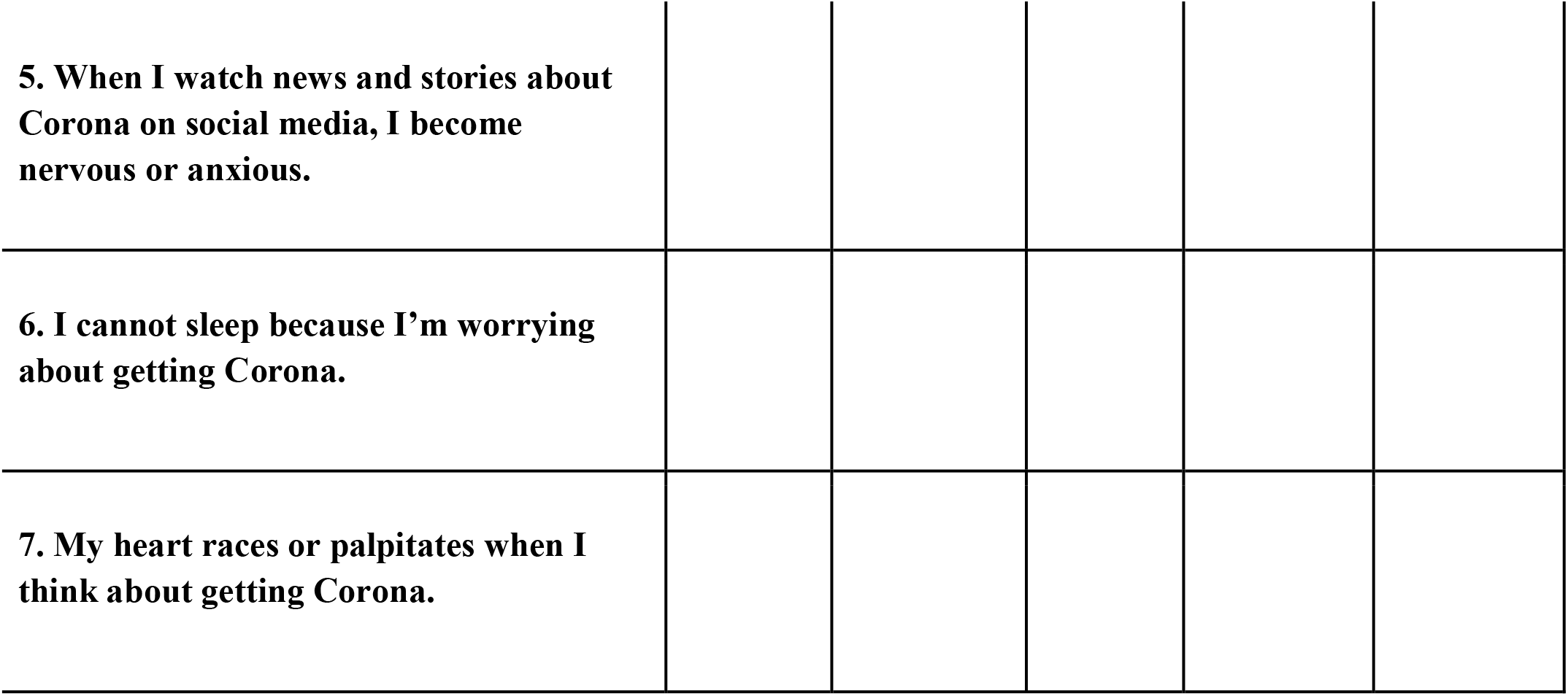

